# Mathematical framework to model Covid-19 daily deaths

**DOI:** 10.1101/2020.05.18.20106104

**Authors:** Poulami Barman, Nabarun Deb, Sumit Mukherjee

## Abstract

The novel coronavirus (2019-nCoV) pandemic has caused widespread socio-economic disruption and, as of 04/07/2020, resulted in more than 72,614 confirmed deaths worldwide. Robust prediction of the trajectory of the death incidence curve is helpful for policy decisions during this ongoing crisis. We propose a non-parametric model to fit the number of daily deaths in a region, which hypothesizes that the death incidence curve will have a convex shape in the beginning, a concave shape near the peak, and a convex shape in the final stage of the death incidence curve after the peak. Using this, we performed robust short-term predictions on phases in five countries worldwide and five US states. Our analysis shows while the five states are all at peaks or past their peaks, US as a country is possibly not at peak yet. Our model can be easily fitted on daily death data from any region.

## Introduction

The novel coronavirus (2019-nCoV) first originated in December 2019 in Wuhan, Hubei province of China, caused by SARS-CoV-2 virus. As of April 7^th^, 2020, World Health Organization has reported 72,614 deaths [1] and about 1.2 million SARS-CoV-2 positive cases worldwide. SARS-CoV-2 virus has mainly three modes of transmission, respiratory transmission, aerosol transmission, and contact transmission[2]. Worldwide travel due to globalization has also resulted in an increased risk of transmission[3]. Starting from the Hubei province, the virus has gradually spread over 185 countries[4] and is still spreading. Between December 2019 and March, 2020, China reported a total of 82,545[5] positive cases, whereas in Italy the number of cases started rising from February 2020[6]. The trends of confirmed cases[4] show the timeline and peak of infection and deaths is variable for each country.

In the absence of a vaccine or a cure, slowing the rates of infection and preparing the healthcare system has been the primary focus for the government across the world. Mathematical modelling has been at the forefront of policy and decision-making to determine timelines for social distancing, stay at home orders, healthcare policies, and city wide lock-downs[7, 8]. A notable amount of research is currently focusing on prediction of cumulative incidence (CI)[9, 10], case fatality rate (CFR) [11-13] and transmission of the disease [14, 15]. Epidemiological models such as Susceptible, Infected and Recovered (SIR), Susceptible, Exposed, Infectious, and Removed (SEIR) have been used to study the transmission based on population mixing [16], travel, reproduction rate and recovery rates [15, 17-20]. An inherent bias in most of these metrics is their dependence on true positive cases, which is hard to estimate during an epidemic [14, 21, 22]. For example, regions with higher testing rates are more likely to observe higher incidence. Thus a meaningful study of the spread of the disease in a region, or comparison across regions, is non-trivial. In contrast, the number of observed deaths due to infections is more robust to these considerations.

Limited research has been published studying the shape of the death incidence curve (DIC) of 2019-nCoV. A recent article by the Institute of Health Metrics and Evaluation (IHME) reported USA’s number of positive cases to peak during first two weeks of April [23] estimating a death toll of 80,000 in the next four months. The same group released a news article projecting that European countries will see their daily death peak during the third week of April[24]. However, both of these death projections have a very wide range of variability (95% CI 38,242–162,106 deaths in US) based on the major assumptions of social distancing in the model. A group of epidemiologists from Imperial College in UK reported a predicted death toll of 550,000 in UK and 2.2million in USA in absence of any social distancing measures[25]. Shortly after that report, they revised their estimate to 5,000 deaths in UK incorporating a perfect social distancing model. The disparity of these predictions highlights the sensitivity of these models on the underlying assumptions. The current global CFR for 2019-nCoV cases is 6.3%, however, Italy’s CFR has been 12.82% whereas in South Korea it has been 2.1% [26]. This degree of variation in the observed CFR across countries is due to the difference in the age distribution, as well as possible overloading of the healthcare system [27]. It remains unclear how situation will progress over time because of dependence on many factors, which is why long term prediction on positive cases or transmission rates remain unreliable. A more tractable question is whether we can understand the short-term trajectory of 2019-nCoV in a robust way to aid in decision making by studying the DIC per region.

## Data Collection

We analyzed two different datasets: a) time-series data of daily 2019-nCoV related deaths for 185 countries and b) Time series data of daily 2019-nCoV related deaths for each USA states. Country-wise time-series data were obtained from John Hopkin’s data platform [4] and state-wise time-series data for daily deaths in USA were obtained from USAfacts.org[28]. For the purpose of testing we have included data through April 12^th^, 2020 for countries and through April 14^th^,2020 for US state level data.

## Methods

To understand the bias in estimation of true positive cases, we have plotted the number of deaths per 1000 infected patients, for five countries.

Motivated by shape of the death incidence curve in China, we consider the 2019-nCoV DIC to be comprised of three functional form segments, a) monotonically increasing convex segment up to an inflection point[29, 30] b) beyond the inflection point a concave function where the curve increases initially to a peak and then decreases to a second inflection point and c) the third segment where the curve is convex and decreases monotonically. We also hypothesize that the peak will not be a single day; instead the peak will be attained across multiple days and thus will look like a plateau (flattened curve).

If we assume that this general pattern will hold for data coming out of a region, we can use this to predict which portion of the DIC a region is currently in. Our proposed model is non-parametric as it only requires a shape restriction on the function in different segments. Thus the model is flexible and gives a good fit to data from different regions. One limitation of our non-parametric model is it’s low predictive power, in that, it will only give reliable short term trend predictions (7days). A parametric model on the other hand can make long term predictions but may fail to adjust to unknown future events such as adherence to the strict lockdown measures, weather, viral mutation and other unknown factors. Thus our model is robust to capture short term trends in a reliable manner, and adapts to sudden changes.

## Formal Methodology in detail

Let [*y*_1_,*y*_2_,..,*y_n_*} denote the observed time-series for number of new deaths indexed by days numbered 1, 2,…. *n*. For each time point *i* ∈ 1,…, *n*, we consider the following three models:

a. Model 1 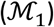

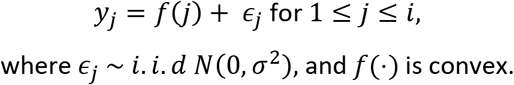
b. Model 2 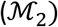

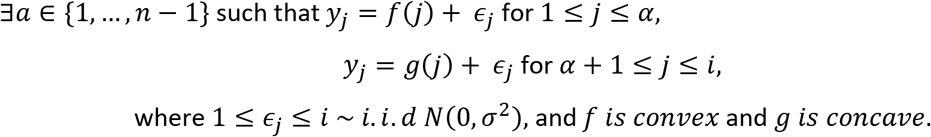
c. Model 3 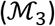

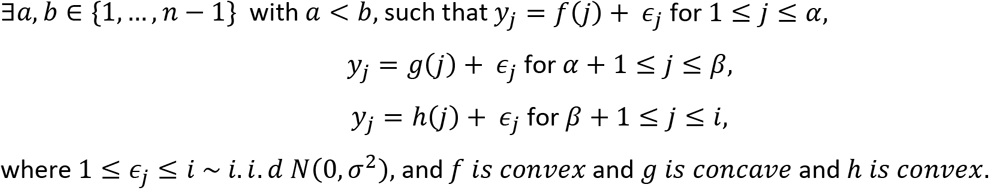

Let *f_i_*(*j*)_1≤_*_j_*_≤_*_i_, g_i_*(*j*)_1≤_*_j_*_≤_*_i_* and *h_i_*(*j*)_1≤_*_j_*_≤_*_i_* denote the fitted values obtained from the data 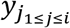 under models 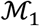, 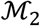 and 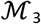 respectively. Then the mean square error (MSE) under 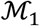, 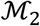, 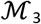 at time point *i* are

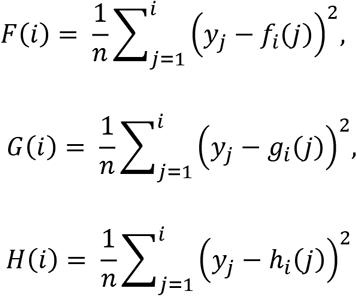

respectively.

Note our models are nested, and so *F*(*i*) ≥ *G*(*i*) ≥ *H*(*i*). In the first segment of the curve (i.e. *i* is a timepoint before the peak) we expect all models 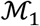, 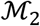, 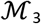 to give the data a good fit, and so we should have *F*(*i*) ≈ *G*(*i*) ≈ *H*(*i*). In the second segment of the curve (*i* is a time point at or near the peak), we expect models 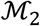 and 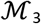 to give a good fit, and 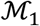 to fail i.e. we should observe *F*(*i*) ≫ *G*(*i*) *≈ H*(*i*). In the final segment of the curve (*i* is a time point after the peak), we expect only 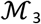 to give a good fit to the data, and so we should have *G*(*i*) ≫ *H*(*i*). Thus if the ratio 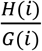 is small, we can say that the peak happens before time point i, and we are in the third segment of the curve. If the ratio 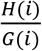 is big, but the ratio 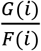 is small, we can say time point *i* is at or near the peak, i.e. we are in the second segment of the curve. And if both the ratios are big, we can say that time point is before the peak, i.e. we haven’t reached the peak and we are in the first segment of the curve. Thus by studying the two ratios 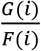 and 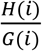 together at a particular i, we can have an understanding of where we lie on the curve. To combine the information across different time points, we compute the above ratios for every time point *i* between 1 and *n*, and study its change with respect to the time variable *i*.

Model 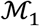 has no inflection points, and can be fitted directly to get the fit, and hence the MSE *F*(*i*). For fitting model 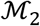 (which has one inflection point *α*), we search over all choices of *α* and fit a convex function to {*y*_1_*, y*_2_*,.., y_α_*}, and a concave function to {*y_α+_*_1_*, y_α+_*_2_*,.., y_i_*}. For every choice of a, we thus get a fitted vector, and a corresponding MSE. We then take a minimum of all the MSE over all possible values of *α* to get the value *G*(*i*). Similarly, for fitting model 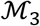, we search over all possible pairs of inflection points(*α*, *β*), and fit a convex function to the data {*y*_1_*, y*_2_*,.., y_α_*}, a concave function to the data {*y_α+_*_1_*, y_α+_*_2_*,.., y_β_*}, and a convex function to the data {*y_β+_*_1_*, y_β+_*_2_*,.., y_i_*}. Having obtained the fit for every value of (*α*, *β*), we again compute the MSE for each such fit, and take a minimum over all possible choices of (*α*, *β*) to get the value *H*(*i*). Before plotting the ratio curves, we smooth out the curves using a cubic spline (penalty 0.6). All calculations were performed using R 3.6.2.

One advantage of our approach is the model can plug in new data each day without needing the recompute previous days’ estimates. We compute the ratios starting from 15 days after the first death in every region because the ratios can be unreliable when computed on initial few days. To see why, note that if there are only four data points, then we can always fit the model 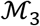 perfectly to give a MSE of 0, as there is always a convex-concave-convex function passing through any four points in the plane. Thus on the 4th day, H(i) = 0 and G(i) can be positive (and small), but the ratio 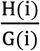 will equal 0, suggesting that the peak is over at day 4, irrespective of the nature of the DIC, which is clearly not a valid conclusion. Similar phenomenon tends to persist for small number of data points, and so the conclusions are reliable only when the ratio 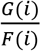 reaches close to 1, after starting from 1 initially and dropping a bit. This suggests that the model 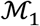 is a good fit, and we have entered the timeline where the data has started showing convexity to a reasonable degree, and not just giving a fit because the number of points is too low.

As mentioned above, both the ratios 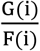 and 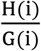 are always less than 1, and we need to come up with reasonable cutoffs to answer the question “how large is too large” in a systematic way, for each of the two curve plots. Specifically, we need to penalize models with increasing complexity, to avoid overfitting. Overfitting in our case would cause curves fitting to our most complex model M3. To account for this, we use a cut off of 0.5 for the curve of 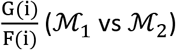, and 0.66 for the curve of 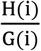 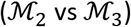. A heuristic behind the choice of the cut offs is that model 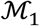 has one choice of a function, and model 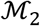 has two choices of a function, so in some sense the ratio of degrees of freedom of the two models should be ½=0.5. Similarly, the heuristic ratio of the degrees of freedom of models 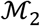 vs 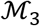 is 2/3=0. 66. These cutoffs will help us distinguish a real dip from a false dip in the two curves 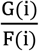 and 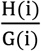 when the case fatality is highly variable. We would consider a dip in 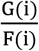 a true one if it drops to its cutoff of 0.5 and continues dropping. Similarly, a dip in 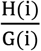 ratio to 0.66 and below is suggestive of a true dip. Any other fluctuations in either of the curves are suggestive of mini peaks.

In the event of a second wave of the DIC, our model can be generalized easily to allow multiple peaks. Assuming that a second peak has the same functional shape, we can have two more models 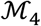 and 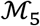, where the models under 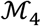 and 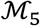 have four segments (convex-concave-convex-concave) and five segments (convex-concave-convex-concave-convex) respectively. Comparing the models 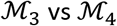 and 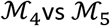, we can predict whether the second peak is approaching, or whether we have crossed the second peak, respectively. Thus, this general schema gives a flexible way to accommodate multiple peaks in the DIC, and can even be trained to fit data with an unknown number of peaks by adaptively estimating the number of peaks.

## Results

For the purpose of the manuscript, we will describe results from five countries in the world: China, Italy, Germany, South Korea and United States of America (USA).

Figure 1 depicts a timeline from January 27^th^ to April 12^th^, 2020. On January 27^th^, China reported a DTT (death toll per thousand positive cases) of 11.35, and by February 16^th^, the DTT decreased to 1.83. South Korea reported a DTT of 3.89 on February 24^th^ and dropped to 0.73 by March 7^th^. USA reported a DTT of 20.20 on March 7^th^ and dropped to 5.01 March 11^th^, 2020. Thus USA started with a higher DTT compared to China and South Korea, and then had a steeper drop. Italy started with a DTT of 14.70 on February 24^th^ dropped to 11 on March 3^rd^ and then rose up to 15.91 by March 15^th^ after which dropped gradually. Germany started the last among the five countries with a DTT of 0.70 on March 11^th^, rose up to 2.05 by April 4^th^ and dropped to 1.89 after that. Italy and Germany both showed a trend of death toll rising after an initial drop in the curve. The initial drop in all countries could be partially accounted to increase in number of testing in each country. Data [31] show that on March 7^th^ USA reported 0.011 total tests being performed per thousand people, and by March 15^th^ it was 0.118 tests per thousand, which was approximately ten-fold increase in testing. By February 24^th^ South Korea was performing 0.558 tests per thousand people, and by March 7^th^ 2020, that increased to 3.476 tests, a 6.22-fold increase. This partially explains the steeper drop in death tolls in US compared to South Korea. Italy reported approximately 5.9-fold increase in testing between February 24^th^ and March 3^rd^. By March 15^th^, even though Italy increased the number of testing by 28 folds, we see an increase in DTT. This increase could be attributed by overload of their healthcare system[32]. Thus from Figure 1, we learn that DTT can vary depending on various factors such as overload of healthcare system, number of testing performed, and others. This observation justifies our consideration to study daily deaths as compared to number of observed positive cases, or CFR (proportional to DTT).

**Figure 1:**
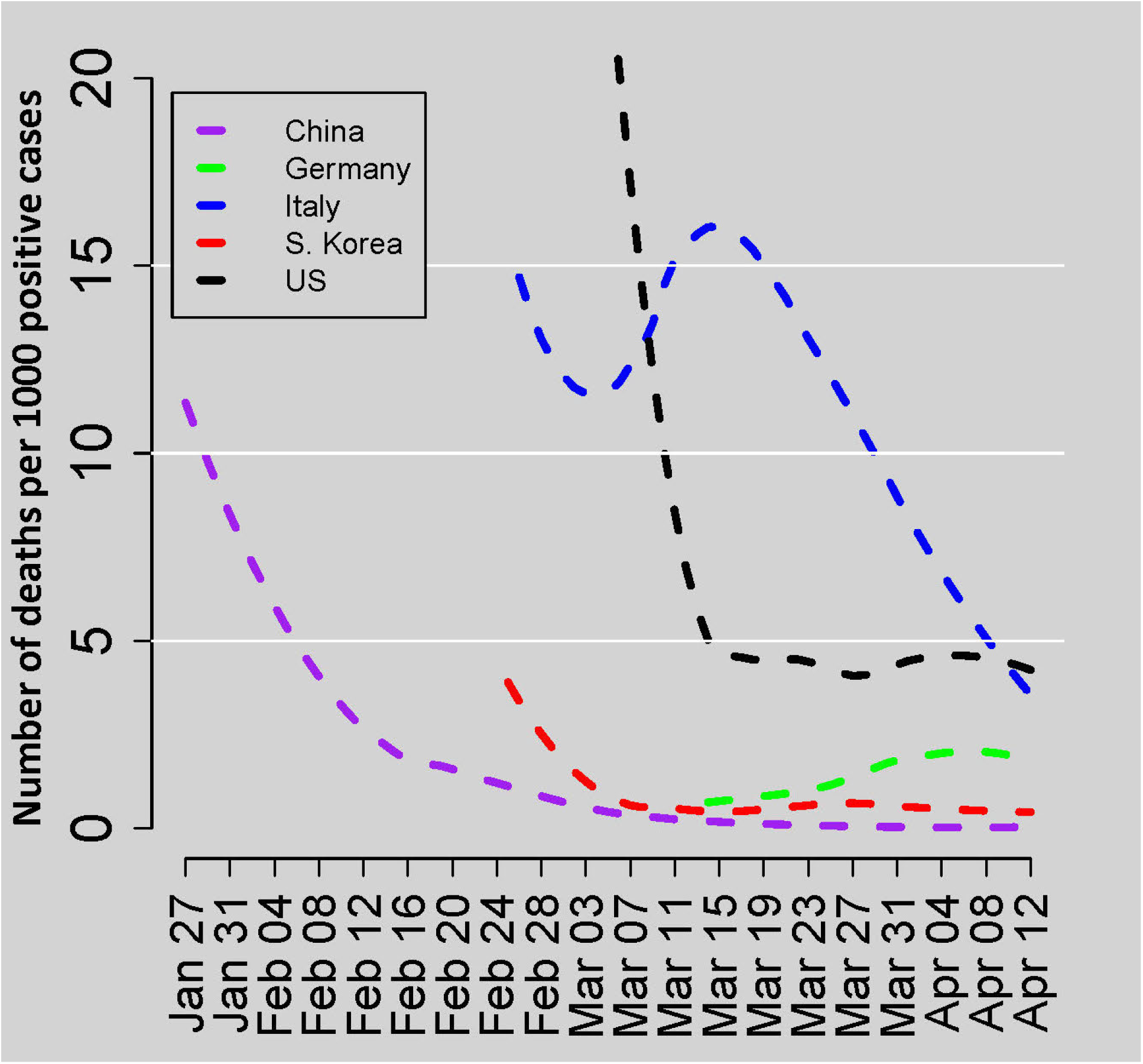
**Plot showing the number of deaths per 1000 positive cases in five countries, plotted over time**.

### Country level results

Figure 2A and 2B both show the DIC in China (black triangles). Figure 2A also overlays the three segments of model 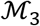 (purple-convex regions, green-concave region). Table 1 reports smoothed values of MSE ratios of 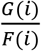 and 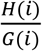 that are plotted in Figure 2C and 2D respectively from China. To see when the peak of the DIC begins, we study Figure 2C, the ratio of MSEs between 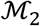 and 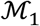, which increases to a peak, and then starts falling sharply around February 17^th^, suggesting that the peak of the DIC began around February 17^th^. Since the curve reached below 0.5, we can also confirm that it is the start of a real peak. Table 2 shows a similar pattern, that around February 17^th^ the ratio 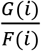 starts to drop sharply, and corresponding ratio of 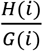 increases sharply. In Figure 2D, we see that the curve 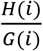 increases to a peak first, and then starts decreasing after March 6^th^, suggesting that the fit for 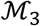 is increasingly better than 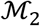 i.e. peak of the DIC is over around March 5^th^. The lowest point in Fig 2D is below 0.66 suggesting that the true peak is over. Thus, China is past the first peak in its DIC.

**Figure 2:**
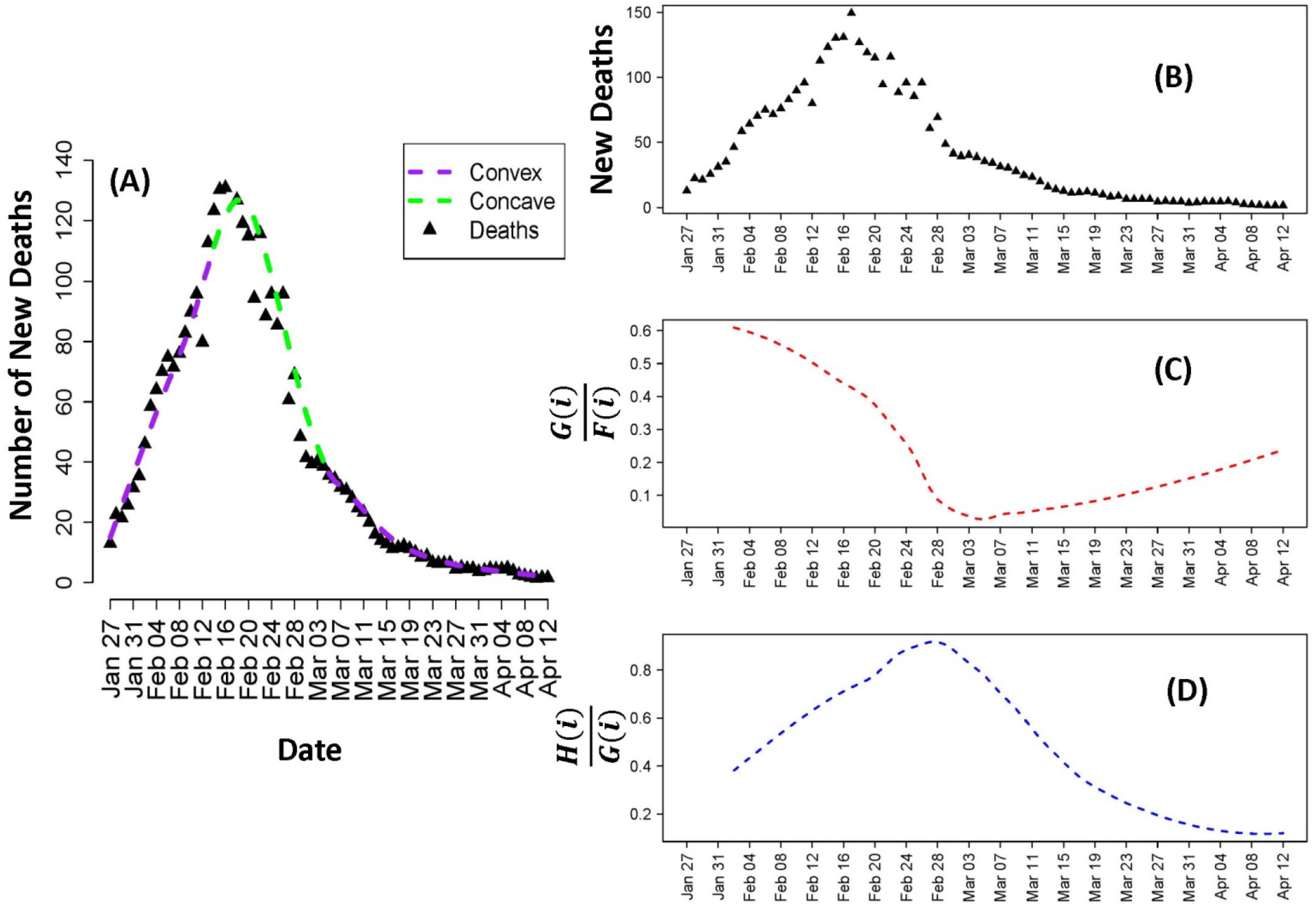
**(A) shows the DIC of China plotting the daily number of deaths over time (black triangles); superimposed purple and green lines show the possible fitted convex and concave portions of the DIC. (B) Daily new deaths have been plotted over time for China (C) Plot showing the ratio of MSE from 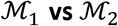 (red curve) and (D) Plot showing ratio of MSE from 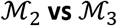 (blue curve)**

**Table 1:** Reports the smoothed ratio of MSEs obtained from, 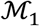, 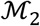 and 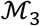 fits in China over time

| Date | Smoothed<br>$\frac{G(i)}{F(i)}$ | Smoothed<br>$\frac{H(i)}{G(i)}$ |
| --- | --- | --- |
| 2-Feb | 0.61 | 0.38 |
| 3-Feb | 0.60 | 0.41 |
| 4-Feb | 0.60 | 0.43 |
| 5-Feb | 0.59 | 0.46 |
| 6-Feb | 0.58 | 0.48 |
| 7-Feb | 0.57 | 0.51 |
| 8-Feb | 0.56 | 0.54 |
| 9-Feb | 0.54 | 0.56 |
| 10-Feb | 0.53 | 0.59 |
| 11-Feb | 0.52 | 0.61 |
| 12-Feb | 0.50 | 0.63 |
| 13-Feb | 0.49 | 0.65 |
| 14-Feb | 0.47 | 0.67 |
| 15-Feb | 0.45 | 0.69 |
| 16-Feb | 0.44 | 0.71 |
| 17-Feb | 0.43 | 0.73 |
| 18-Feb | 0.41 | 0.74 |
| 19-Feb | 0.40 | 0.76 |
| 20-Feb | 0.38 | 0.78 |
| 21-Feb | 0.35 | 0.81 |
| 22-Feb | 0.32 | 0.84 |
| 23-Feb | 0.29 | 0.87 |
| 24-Feb | 0.26 | 0.89 |
| 25-Feb | 0.22 | 0.90 |
| 26-Feb | 0.17 | 0.91 |
| 27-Feb | 0.12 | 0.92 |
| 28-Feb | 0.09 | 0.92 |
| 29-Feb | 0.07 | 0.91 |
| 1-Mar | 0.05 | 0.88 |
| 2-Mar | 0.04 | 0.86 |
| 3-Mar | 0.04 | 0.83 |
| 4-Mar | 0.03 | 0.80 |
| 5-Mar | 0.03 | 0.78 |
| 6-Mar | 0.03 | 0.74 |
| 7-Mar | 0.04 | 0.70 |
| 8-Mar | 0.05 | 0.67 |
| 9-Mar | 0.05 | 0.64 |
| 10-Mar | 0.05 | 0.60 |
| 11-Mar | 0.05 | 0.55 |
| 12-Mar | 0.06 | 0.51 |
| 13-Mar | 0.06 | 0.48 |
| 14-Mar | 0.06 | 0.45 |
| 15-Mar | 0.07 | 0.42 |
| 16-Mar | 0.07 | 0.38 |
| 17-Mar | 0.07 | 0.36 |
| 18-Mar | 0.08 | 0.33 |
| 19-Mar | 0.08 | 0.31 |
| 20-Mar | 0.09 | 0.29 |
| 21-Mar | 0.09 | 0.28 |
| 22-Mar | 0.10 | 0.26 |
| 23-Mar | 0.10 | 0.25 |
| 24-Mar | 0.11 | 0.23 |
| 25-Mar | 0.11 | 0.22 |
| 26-Mar | 0.12 | 0.21 |
| 27-Mar | 0.13 | 0.19 |
| 28-Mar | 0.13 | 0.18 |
| 29-Mar | 0.14 | 0.17 |
| 30-Mar | 0.15 | 0.16 |
| 31-Mar | 0.15 | 0.16 |
| 1-Apr | 0.16 | 0.15 |
| 2-Apr | 0.16 | 0.14 |
| 3-Apr | 0.17 | 0.13 |
| 4-Apr | 0.18 | 0.13 |
| 5-Apr | 0.19 | 0.13 |
| 6-Apr | 0.19 | 0.12 |
| 7-Apr | 0.20 | 0.12 |
| 8-Apr | 0.21 | 0.12 |
| 9-Apr | 0.21 | 0.12 |
| 10-Apr | 0.22 | 0.12 |
| 11-Apr | 0.23 | 0.12 |
| 12-Apr | 0.24 | 0.12 |

Using the same principles outlined above, we also fitted death data from France, Germany, Italy, South Korea and USA (Figure 3). Figure 3 reports ratio of 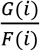 and 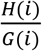 for four countries in each of its quadrant (A, B, C, D). Quadrant A shows 3 curves for Germany. The first plot in quadrant A show the number daily new deaths over time. The second plot (red curve) in quadrant A plots 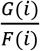 over time and the third plot shows 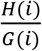 over time for Germany. Quadrant B, C and D show the same series of curves a) number of daily new deaths over time b) 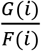 over time and c) 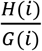 over time for Italy, South Korea and USA respectively. Studying the curves, we have listed an approximate peak start and peak end dates in Table 2. Germany has started its peak from April 2^nd^, and is still at peak. Italy has started its peak around March 24^th^ and escaped peak on April 10^th^ (Fig 3B). Based on the slope of the trend of 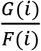 and 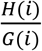 values (Fig 3B), we can also get a sense of how soon the peak will be over. South Korea (Fig 3C) had a first peak from March 5^th^ - March 18^th^, and is going through a second peak from April 6^th^. USA has reached a mini peak starting April 1^st^ (Fig 3D), but there is no sharp fall, so it is unclear whether this is the main peak or a mini peak. In summary, China, Italy and South Korea have crossed their first peaks and South Korea and China are starting their second peaks. The smoothed ratios of 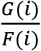 and 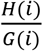 for each country are presented in Supplemental tables (ST1).

**Figure 3:**
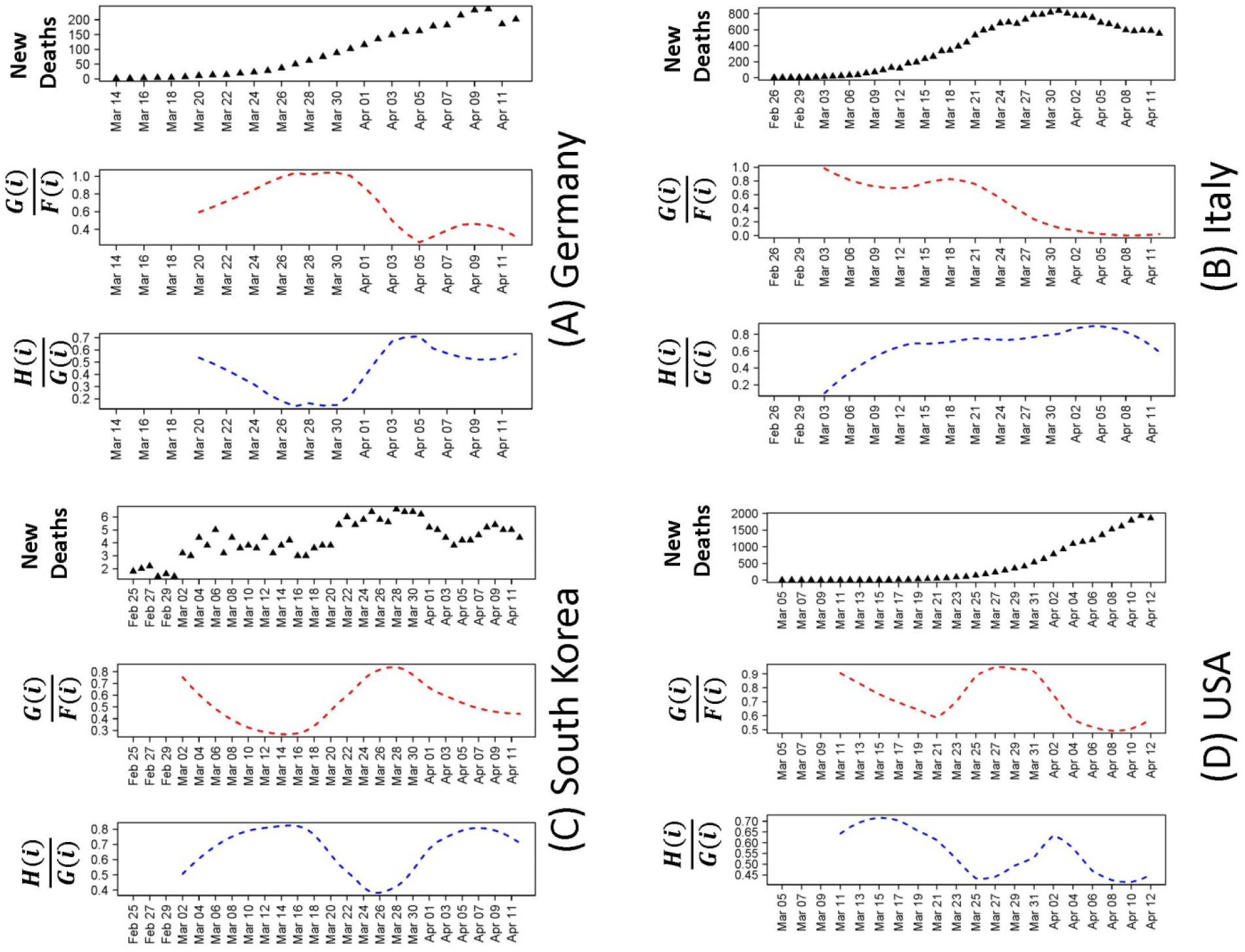
*(A) Germany: Scatter Plot of number of new deaths per day, ratio of MSE from* 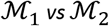 *and ratio of MSE from* 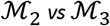 *(B) Italy: Scatter Plot of number of new deaths per day, ratio of MSE from* 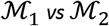 *and ratio of MSE from* 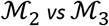. *(C) South Korea: Scatter Plot of number of new deaths per day, ratio of MSE from* 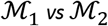 *and ratio of MSE from* 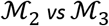. *(D) USA: Scatter Plot of number of new deaths per day, ratio of MSE from* 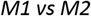 *and ratio of MSE from* 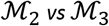.

**Table 2:** Table showing the estimated start and end dates for peak by each of the five countries, China, Germany, Italy, South Korea and USA. Blank cells indicate that the appropriate date is not reached

| Country | Begin Peak | End Peak | Comments |
| --- | --- | --- | --- |
| China | February 9<sup>th</sup> | March 5<sup>th</sup> | Passed one peak, slowly approaching second one |
| Germany | April 2<sup>nd</sup> | At peak | Peak starting at April 2<sup>nd</sup>, still at peak. |
| Italy | March 24<sup>th</sup> | April 10<sup>th</sup> | Peak over, entering third stage |
| South Korea | i) March 5<sup>th</sup><br>ii) April 6<sup>th</sup> | i) March 18<sup>th</sup><br>ii) At peak | First peak from 5<sup>th</sup> March-18<sup>th</sup> March, second peak from 6<sup>th</sup> April, exiting peak soon. |
| United States of America |  | -- | Has reached a mini peak starting April 1 |

### US State level results

Within USA, we report results from five states: New York (NY), Washington (WA), California (CA), Michigan (MI) and New Jersey (NJ). Figure 4 shows the plots of the ratios of 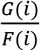 and 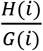 of NY and WA. The smoothed ratios of 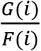 and 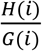 for each country are presented in Supplemental tables (ST2).

Figure 4A shows the series of three plots for NY; the number of daily new deaths (Fig 4A-a), ratio of 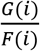 over time (Fig 4A-b) and 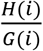 over time (Fig 4A-c). The red curve (Fig 4 A-b) has started a dip around April 8^th^ suggesting NY has entered its peak, and the blue curve (Fig 4 A-c) shows that NY is still in peak. Figure 4B shows the series of three plots for state of WA; the number of daily new deaths (Fig 4B-a), ratio of 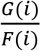 over time (Fig 4B-b) and 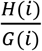 over time (Fig 4B-c). Figure 4B-b shows the red curve starts the first dip around March 17^th^showing a start of a mini peak. Figure 4B-c shows start of a dip around March 21^st^, suggesting end of the mini peak. Fig 4B-b shows a second dip around April 8^th^ and keeps dropping indicating that the second peak has begun in WA, and figure 4B-c shows that the peak is still continuing. The plots showing the daily deaths over time, 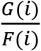 and 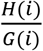 over time for NJ, MI and CA are available in supplemental figures (S1, S2, S3). CA and NJ have started their peaks around April 8^th^, and 4^th^ respectively, and are still in peak (Table 3). Michigan has completed its first peak from April 2^nd^ to April 6^th^ and has started a second peak on April 11^th^ and still in peak. Table 3 contains the list of peak start and end dates for all five US states.

**Figure 4:**
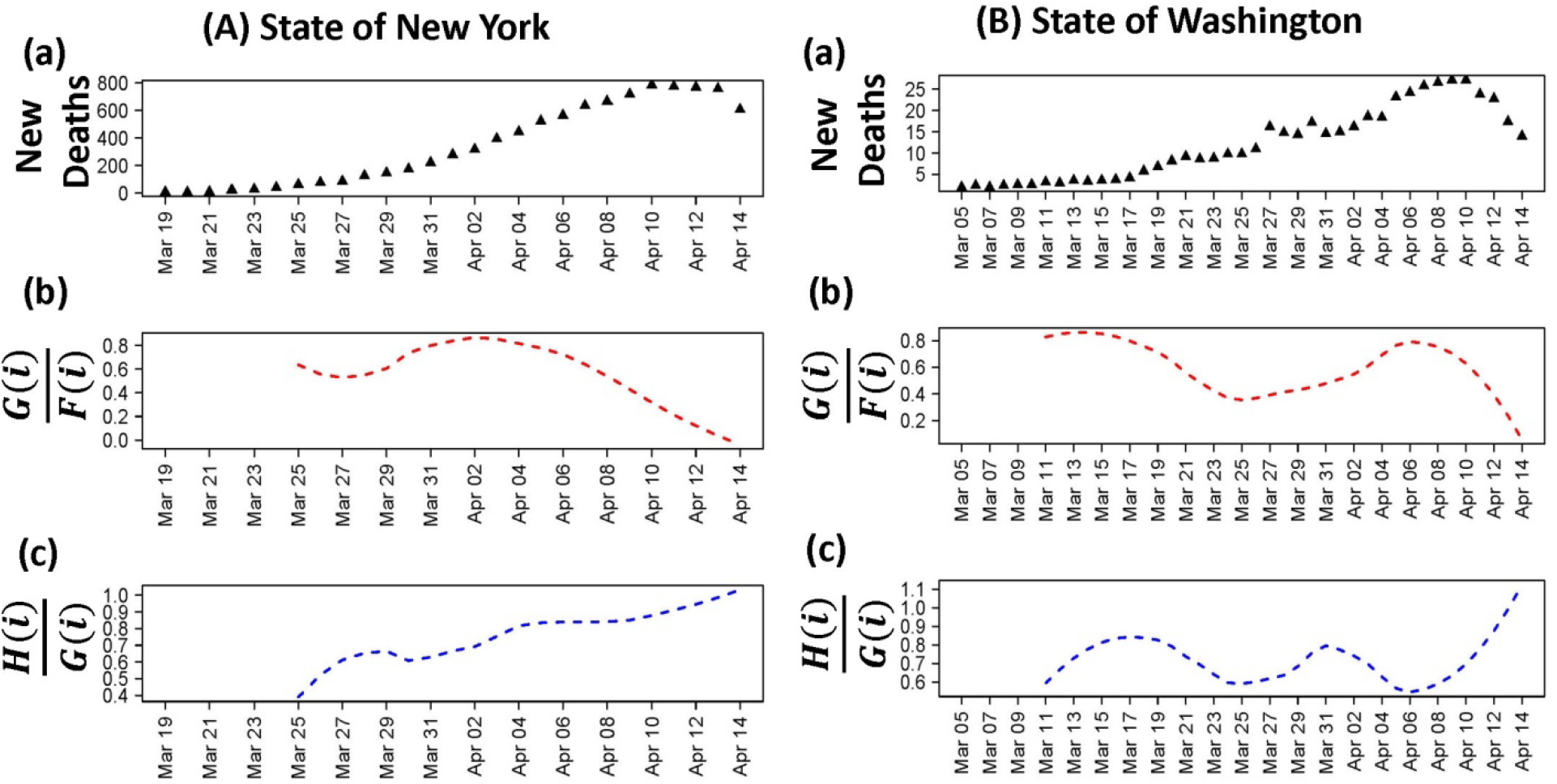
*(A) State of New York: Scatter Plot of number of (a) new deaths per day, (b) ratio of MSE from* 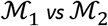 *and (c) ratio of MSE from* 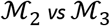 *(B) State of Washington: Scatter Plot of number of (a) new deaths per day, (b)ratio of MSE from M1 vs M2 and (c)ratio of MSE from* 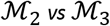.

**Table 3:** Table showing the estimated start and end dates for peak by each USA states

| US States | Begin Peak | End Peak | Comments |
| --- | --- | --- | --- |
| New York | April 8<sup>th</sup> | At peak | At peak |
| Washington | April 8<sup>th</sup> | At peak | Completed a mini peak from March 17<sup>th</sup> -21<sup>st</sup>. At peak. |
| California | April 8<sup>th</sup> | At peak | At peak |
| Michigan | i) April 2<sup>nd</sup><br>ii) April 11<sup>th</sup> | i) April 6<sup>th</sup><br>ii) At peak | First peak between April 2<sup>nd</sup> – April 6<sup>th</sup>. Started a second peak on April 6<sup>th</sup> and still in peak |
| New Jersey | April 4<sup>th</sup> | At peak | At peak |

## Discussion

This study was predicated with the aim of providing a generalized mathematical framework to model DIC of a region, irrespective of the extent of 2019-nCoV spread and population density of the region. Currently, the worldwide pandemic of 2019-nCoV is primarily managed by various state and nationally issued policies involving social distancing and lockdowns. The goal of the study was to robustly and accurately model the current trend in the DIC and identify peaks i.e. when the rate of deaths starts to flatten. This is essential to guide control efforts and to plan health care system requirements.

We used non-parametric shape-restricted regression [33, 34] (convex-concave-convex) to fit the number of new deaths across time. Our model is able to predict at what stage a region is in its 2019-nCoV timeline, from about a week ahead. Our non-parametric approach gives a robust fit despite of considerable variability in the number of cases on each day. In the study we have identified start dates of peaks in five countries and five US states. Our model can be easily be fitted on data from other regions. USA has reached a peak, but most likely this is a mini peak. Our model suggests that there is a trend of a second peak in Asian countries such as South Korea and China, which is also validated by a recent study [35]. Italy has crossed its peak, i.e. case fatality in Italy has started declining. Within USA, some of the states have already reached their peaks. California as a state has been able to better contain the virus, as it had an early start and has already reached its peak, but has a lower total number of deaths per million compared to some of the other states (NY and NJ). Overall, since USA is at a mini peak, it seems that lockdown is working to slow down the rates of 2019-nCoV spread. However, since Asian countries are approaching a second one[35], it is reasonable to think, for USA, we anticipate an increase in the number of deaths in the coming months.

As indicated before, our model, while providing robust guidelines to what may happen in the near future, fails to predict long term behavior of the DIC, as well as cumulative deaths. In the absence of widespread serological tests, currently there is debate on the percentage of asymptomatic cases in the population. Studies have reported vastly different asymptomatic cases, ranging from 51.4% [36] in Diamond princess ship and 87.9% [37] among pregnant women in New York. Variability in percentage of asymptomatic cases, long incubation period of the virus coupled with current testing on only symptomatic cases, incorporate bias in estimation of either the number of positive cases. On the other hand, there are studies [18] claiming that CFR underestimates the true mortality rate in the population due to the infection. Our analysis re-emphasizes that CFR is affected by multiple unknown factors such as disease prevalence[38], testing rates, right censoring[18], human behavior, that are challenging to account for. Thus, we chose number of daily new deaths as our statistic, and relied on a non-parametric model to predict short term behavior. Currently the existing predictions based on parametric modelling assumptions [25] produce estimates with a wide range of uncertainty, and the estimates change considerably over the span of few days, which can be problematic for policy decisions.

In conclusion, we believe our model will be beneficial to identify changing trends. Being able to predict the start and end of peak in DIC in a region is important for policy makers worldwide. This will enable them to mobilize resources (such as, equipment, testing kits, health care professionals) as needed. Since our model can be used to assess peaks in any region, it can be adapted easily to model country, state or county level data. A prolonged national lockdown can have a negative impact both for small and large businesses, leading to a financial crisis. In the efforts of reopening economy, our model can help to identify low-risk regions which are already past their peak.

## Data Availability

We analyzed two different publicly available datasets: a) time-series data of daily 2019-nCoV related deaths for 185 countries and b) Time series data of daily 2019-nCoV related deaths for each USA states. Country-wise time-series data were obtained from John Hopkin's data platform and state-wise time-series data for daily deaths in USA were obtained from USAfacts.org. For the purpose of testing we have included data through April 12th, 2020 for countries and through April 14th,2020 for US state level data.

https://usafacts.org/visualizations/coronavirus-covid-19-spread-map/

https://github.com/CSSEGISandData/COVID-19

## Conflict of interest statement

None

## Financial support

^#^Research partially supported by NSF Grant DMS-1712037

## Notes

### Competing Interest Statement

The authors have declared no competing interest.

### Funding Statement

Research partially supported by NSF Grant DMS-1712037. No third party payments were received by any institution or any of the authors for this project at any time.

## Reference

1. Organization, W.H. Novel Coronavirus (COVID-19) Situation Report – 78. 2020 April 7; Available from: https://www.who.int/docs/default-source/coronaviruse/situation-reports/20200407-sitrep-78-covid-19.pdf?sfvrsn=bc43e1b2.

2. Rothan, H.A. and S.N. Byrareddy, The epidemiology and pathogenesis of coronavirus disease (COVID-19) outbreak. Journal of autoimmunity, 2020: p. 102433.

3. Mack, A., E.R. Choffnes, and D.A. Relman, Infectious disease movement in a borderless world: workshop summary. 2010: National Academies Press.

4. Dong, E., H. Du, and L. Gardner, An interactive web-based dashboard to track COVID-19 in real time. The Lancet infectious diseases, 2020.

5. Organization, W.H. Coronavirus disease 2019 (COVID-19) Situation Report—71. 2020 March 31; Available from: https://www.who.int/docs/default-source/coronaviruse/situation-reports/20200331-sitrep-71-covid-19.pdf?sfvrsn=4360e92b8.

6. Organization, W.H. Novel Coronavirus (COVID-19) Situation Report – 19. 2020 February 8; Available from: https://www.who.int/docs/default-source/coronaviruse/situation-reports/20200208-sitrep-19-ncov.pdf?sfvrsn=6e091ce64.

7. Prem, K., et al., The effect of control strategies to reduce social mixing on outcomes of the COVID-19 epidemic in Wuhan, China: a modelling study. The Lancet Public Health, 2020.

8. De Brouwer, E., D. Raimondi, and Y. Moreau, Modeling the COVID-19 outbreaks and the effectiveness of the containment measures adopted across countries. medRxiv, 2020: p. 2020.04.02.20046375.

9. Sebastiani, G., M. Massa, and E. Riboli, Covid-19 epidemic in Italy: evolution, projections and impact of government measures. European Journal of Epidemiology, 2020: p. 1.

10. Pham, H., On Estimating the Number of Deaths Related to Covid-19. Mathematics, 2020. 8(5): p. 655.

11. Onder, G., G. Rezza, and S. Brusaferro, Case-fatality rate and characteristics of patients dying in relation to COVID-19 in Italy. Jama, 2020.

12. Rajgor, D.D., et al., The many estimates of the COVID-19 case fatality rate. The Lancet Infectious Diseases, 2020.

13. Mahase, E., Coronavirus: covid-19 has killed more people than SARS and MERS combined, despite lower case fatality rate. 2020, British Medical Journal Publishing Group.

14. Battegay, M., et al., 2019-novel Coronavirus (2019-nCoV): estimating the case fatality rate–a word of caution. Swiss medical weekly, 2020. 150(0506).

15. Kucharski, A.J., et al., Early dynamics of transmission and control of COVID-19: a mathematical modelling study. The lancet infectious diseases, 2020.

16. Law, G.R., et al., What do epidemiologists mean by ‘population mixing’? Pediatric blood & cancer, 2008. 51(2): p. 155–160.

17. Lopez, L.R. and X. Rodo, A modified SEIR model to predict the COVID-19 outbreak in Spain and Italy: simulating control scenarios and multi-scale epidemics. medRxiv, 2020: p. 2020.03.27.20045005.

18. Hauser, A., et al., Estimation of SARS-CoV-2 mortality during the early stages of an epidemic: a modelling study in Hubei, China and northern Italy. medRxiv, 2020: p. 2020.03.04.20031104.

19. Wang, H., et al., Phase-adjusted estimation of the number of Coronavirus Disease 2019 cases in Wuhan, China. Cell Discovery, 2020. 6(1): p. 1–8.

20. Wu, J.T., K. Leung, and G.M. Leung, Nowcasting and forecasting the potential domestic and international spread of the 2019-nCoV outbreak originating in Wuhan, China: a modelling study. The Lancet, 2020. 395(10225): p. 689–697.

21. Lipsitch, M., et al., Potential biases in estimating absolute and relative case-fatality risks during outbreaks. PLoS neglected tropical diseases, 2015. 9(7).

22. Wang, C., et al., A novel coronavirus outbreak of global health concern. The Lancet, 2020. 395(10223): p. 470–473.

23. COVID, I. and C.J. Murray, Forecasting COVID-19 impact on hospital bed-days, ICU-days, ventilator-days and deaths by US state in the next 4 months. medRxiv, 2020.

24. Evaluation, I.o.H.M.a., New COVID-19 forecasts for Europe: Italy & Spain have passed the peak of their epidemics; UK, early in its epidemic, faces a fast-mounting death toll. 2020, Institute of Health Metrics and Evaluation.

25. Neil M Ferguson, D.L., Gemma Nedjati-Gilani, Natsuko Imai, Kylie Ainslie, Marc Baguelin,, et al. Report 9: Impact of non-pharmaceutical interventions (NPIs) to reduce COVID-19 mortality and healthcare demand. March 16, 2020; Available from: https://spiral.imperial.ac.uk:8443/bitstream/10044/1/77482/8/2020-03-16-COVID19-Report-9.pdf.

26. Organization, W.H. Coronavirus disease 2019 (COVID-19) Situation Report—85. 2020 April 14; Available from: https://www.who.int/docs/default-source/coronaviruse/situation-reports/20200414-sitrep-85-covid-19.pdf?sfvrsn=7b8629bb4.

27. Dowd, J.B., et al., Demographic science aids in understanding the spread and fatality rates of COVID-19. medRxiv, 2020: p. 2020.03.15.20036293.

28. USAfacts.org. Coronavirus Locations: COVID-19 Map by County and State. 2020 [cited 2020; Available from: https://usafacts.org/visualizations/coronavirus-covid-19-spread-map/.

29. Page, E., On problems in which a change in a parameter occurs at an unknown point. Biometrika, 1957. 44(1/2): p. 248–252.

30. Pikard, D., Testing and estimating change-point in time series. Adv. Appl. Probab, 1985. 7: p. 841–867.

31. Joe Hasell, E.O.-O., Edouard Mathieu, Hannah Ritchie, Diana Beltekian, Bobbie Macdonald and Max Roser. Coronavirus (COVID-19) Testing. 2020; Available from: https://ourworldindata.org/grapher/full-list-cumulative-total-tests-per-thousand.

32. Grasselli, G., A. Pesenti, and M. Cecconi, Critical care utilization for the COVID-19 outbreak in Lombardy, Italy: early experience and forecast during an emergency response. Jama, 2020.

33. Samworth, R. and B. Sen, Special Issue on” Nonparametric Inference Under Shape Constraints”. 2018.

34. Groeneboom, P. and G. Jongbloed, Nonparametric estimation under shape constraints. Vol. 38. 2014: Cambridge University Press.

35. Strzelecki, A., The second worldwide wave of interest in coronavirus since the covid-19 outbreaks in south korea, italy and iran: A google trends study. arXiv preprint arXiv:2003.10998, 2020.

36. Mizumoto, K., et al., Estimating the asymptomatic proportion of coronavirus disease 2019 (COVID-19) cases on board the Diamond Princess cruise ship, Yokohama, Japan, 2020. Eurosurveillance, 2020. 25(10): p. 2000180.

37. Desmond Sutton, M.D., Karin Fuchs, M.D., M.H.A., Mary D’Alton, M.D., Dena Goffman, M.D. Universal Screening for SARS-CoV-2 in Women Admitted for Delivery. 2020 April 13, 2020; Available from: https://www.nejm.org/doi/full/10.1056/NEJMc2009316.

38. Bendavid, E., et al., COVID-19 Antibody Seroprevalence in Santa Clara County, California. medRxiv, 2020: p. 2020.04.14.20062463.

